# Factors associated with viral load suppression and indicators of stigma among people living with HIV in Dar es Salaam tertiary hospitals, Tanzania

**DOI:** 10.1101/2022.09.02.22279535

**Authors:** Mary Spicar Kilapilo, Idda Hubert Mosha, George Msema Bwire, Godfrey Leonard Sambayi, Raphael Zozimus Sangeda, Japhet Killewo

**Author notes:** Correspondence author: (RZS).

## Abstract

Perception of stigma can contribute to virological failure among people living with HIV (PLHIV). This study was conducted to assess the effect of stigmatization and self-discrimination on viral load suppression among PLHIV at tertiary hospitals in Dar es Salaam, Tanzania. This was a hospital-based cross-sectional study conducted in Temeke Regional Referral Hospital (RRH) and Amana RRH at the Care and Treatment Clinic (CTC) between July and August 2020 using a structured questionnaire with open and close-ended questions. Factors for stigmatization and viral load suppression were compared using the Chi-square test, while factors for viral load suppression were analyzed using multinomial logistic regression analysis. Altogether, 406 PLHIV participated, with most female respondents, 298 (73.2%). The majority (50%) were aged between 25 and 44 years, whereas 171 (42.5%) respondents were married. Most of the participants, 382 (94.6%), were on a dolutegravir-based regimen, with the majority, 215 (52.8%), having a refilling interval of three months. The majority of the respondents, 379 (93.1%), disclosed their status. Most participants, 355 (87.4%), preferred having a separate HIV clinic, while 130 (32.1%) participants were not ready to be attended by the health care workers (HCWs) familiar to them. Male patients were 60% less likely to suppress their viral load as compared to female patients (adjusted odds ratio [aOR]: 0.4, 95% confidence interval [95%]: 0.19 – 0.77, p = 0.007). Refill interval was significantly associated with viral load suppression. For example, patients with a one-month refill interval had odds of 0.01 (95% CI: 0.003-0.42, p < 0.0001) compared to six-month refill intervals. Stigmatization elements appeared to influence the viral load suppression among PLHIV. Factors such as gender and refill time interval were significantly associated with viral load suppression among HIV patients in the Dar es Salaam region.

## Introduction

Viral load suppression indicates disease progression, wellness, antiretroviral therapy (ART) effectiveness and minimized HIV drug resistance (HIVDR). On the other hand, virological failure indicates the inverse and is associated with HIVDR [1]. Upon characterization of the factors contributing to non-adherence, stigmatization was found to have a major link with non-adherence [2,3].

PLHIV continue to suffer from stigma and discrimination from their family and communities. Evidence from research suggests that HIV-related stigma and self-stigmatization result in a delay in the disclosure of HIV serostatus, which is a potential barrier to HIV counseling, retention in care and treatment, and uptake of and adherence to ART [4]. AIDS-related stigma and discrimination impede millions of PLHIV from accessing and benefiting from effective prevention and treatment services [5].

Self-stigmatization among PLHIV is one of the consequences of stigma from the community. The perception may force PLHIV to hide their serostatus and, in many cases, to continue engaging in high-risk behaviors [6]. Therefore, this study was conducted to assess the effect of stigmatization and self-discrimination on viral load suppression among PLHIV attending care and treatment clinics at Amana and Temeke RRHs in Dar es Salaam, Tanzania.

## Materials and methods

### Study design and area and period of the study

A hospital-based cross-sectional study was conducted in Temeke Regional Referral Hospital (RRH) and Amana RRH at the Care and Treatment Clinic (CTC) in Dar es Salaam, Tanzania, between July and August 2020. Dar es Salaam city was selected to represent the regions found in Tanzanian Mainland. Dar es Salaam is the largest city, business and the former capital of Tanzania, with an approximated population of more than five million people (almost 10% of the country’s population) [7].

### Study population and sampling

A total of 406 PLHIV aged above 18 years and used antiretrovirals (ARVs) for more or equal to six months and attended Amana and Temeke RRHs participated in this study. The sample size was derived using the cross-sectional formula [8] and the assumed prevalence from the previous study [9]. A systematic approach was used during the recruitment of research participants. This was done by obtaining a sampling interval “n” (total patients on CTC divided by 200). Then a patient was sampled after every “n” interval.

### Data collection tool

A structured questionnaire with open and close-ended questions was prepared following an intensive literature review on a topic related to stigmatization, adherence and viral load suppression [9–11]. The English questionnaire was designed and translated to Kiswahili (local language). The Kiswahili questionnaire was uploaded on REDCap (Research Electronic Data Capture). REDCap is an electronic data capture tool hosted at Muhimbili University of Health and Allied Sciences (MUHAS) [12,13]. The questionnaire was tested on a pilot population of 30 patients (15 from each CTC). The updated Kiswahili questionnaire was then used to collect patients’ demographics, pharmacy refill, and ART adherence factors. Data was collected using tablets and coded in the REDcap after being translated into English. REDCap data were downloaded, cleaned in excel (to remove incomplete forms), then exported to Statistical Package for Social Sciences (SPSS software version 25, Chicago Inc., USA) for analysis.

### Data management and analysis

Descriptive statistics were summarized using frequencies and percentages. Stigmatization was estimated using the HIV stigma toolkit described elsewhere [14]. A Chi-square test was employed to analyze the association between categorical variables such as gender, refill interval, regimen used and HCWs preference and viral load suppression. Viral load suppression and high viral load were defined as viral load counts below 1,000 copies/ ml and above 1,000 copies/ ml, respectively [15]. Factors associated with viral load suppression were controlled using a binary regression model. Factors with a p-value less than 0.2 in univariate regression were qualified for multivariate logistic regression [16]. A p-value of less than 0.05 was considered statistically significant.

### Ethical considerations

Ethical clearance was sought from the MUHAS Institutional Review Board (DA.25/111/01/10/02/2021). Temeke RRH and Amana RRH permitted the study to be conducted on their respective premises and patients. All participants provided written informed consent for participation. The consent included information on the description of the study, data privacy/ confidentially and handling. Patients who reported self-discrimination and stigmatization were counseled on medication adherence. Preventive measures for the COVID-19 pandemic were observed during the whole period of data collection.

## Results

Overall, 406 PLHIV were included in this study, with 205 PLHIV from Amana RRH and 201 PLHIV from Temeke RRH. Most of the respondents were female, 298 (73.2%), and the majority (50%) were between 25 and 44 years old. The majority of the respondents, 151 (38.2%), received ART for 7 to 12 years. In addition, the majority of study respondents, 171 (42.5%), were married and most participants, 382 (94.6%), reported to be on a dolutegravir-based regimen. The majority, 215 (52.8%), had a refill interval of three months (Table 1).

**Table 1.**
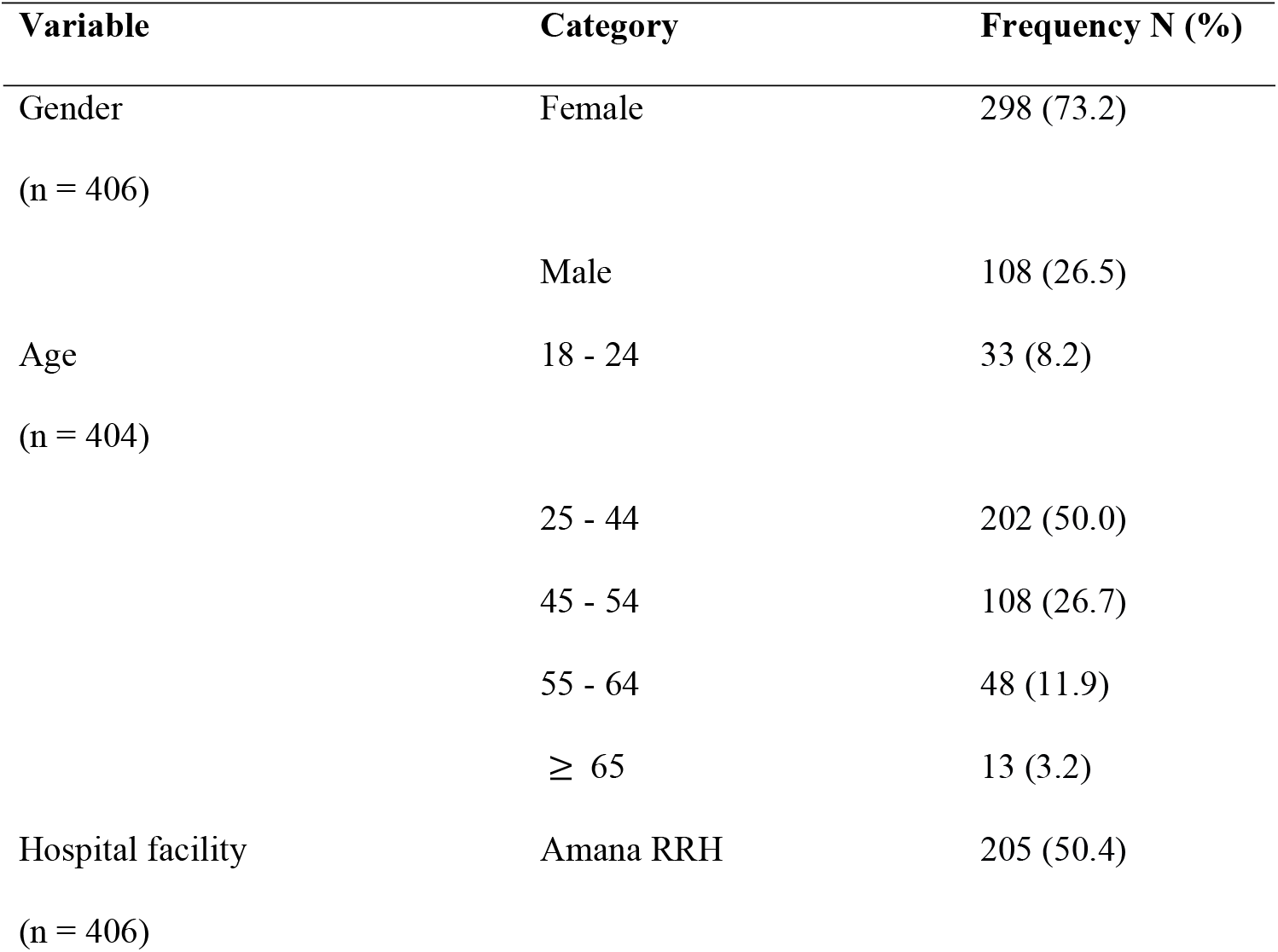
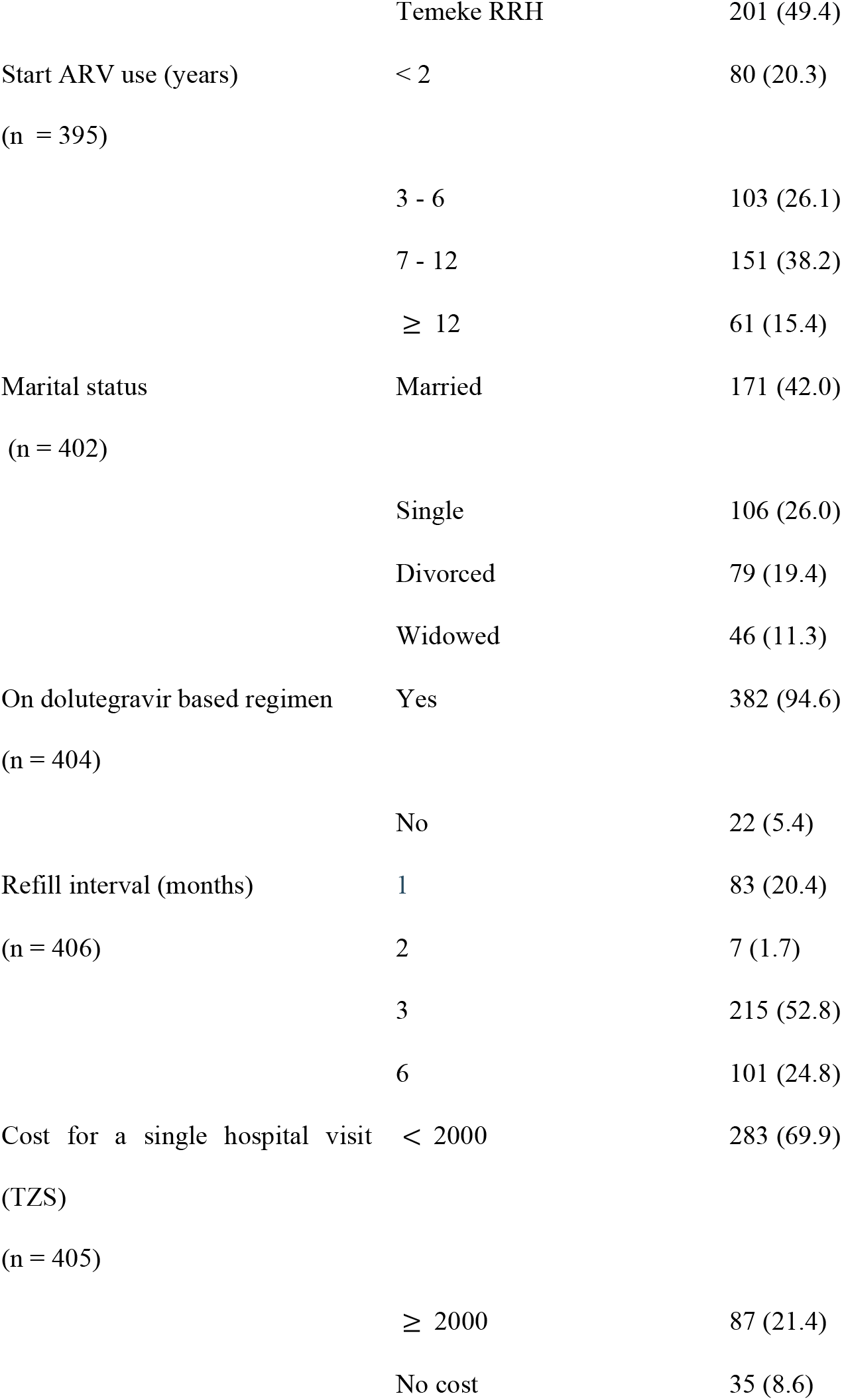
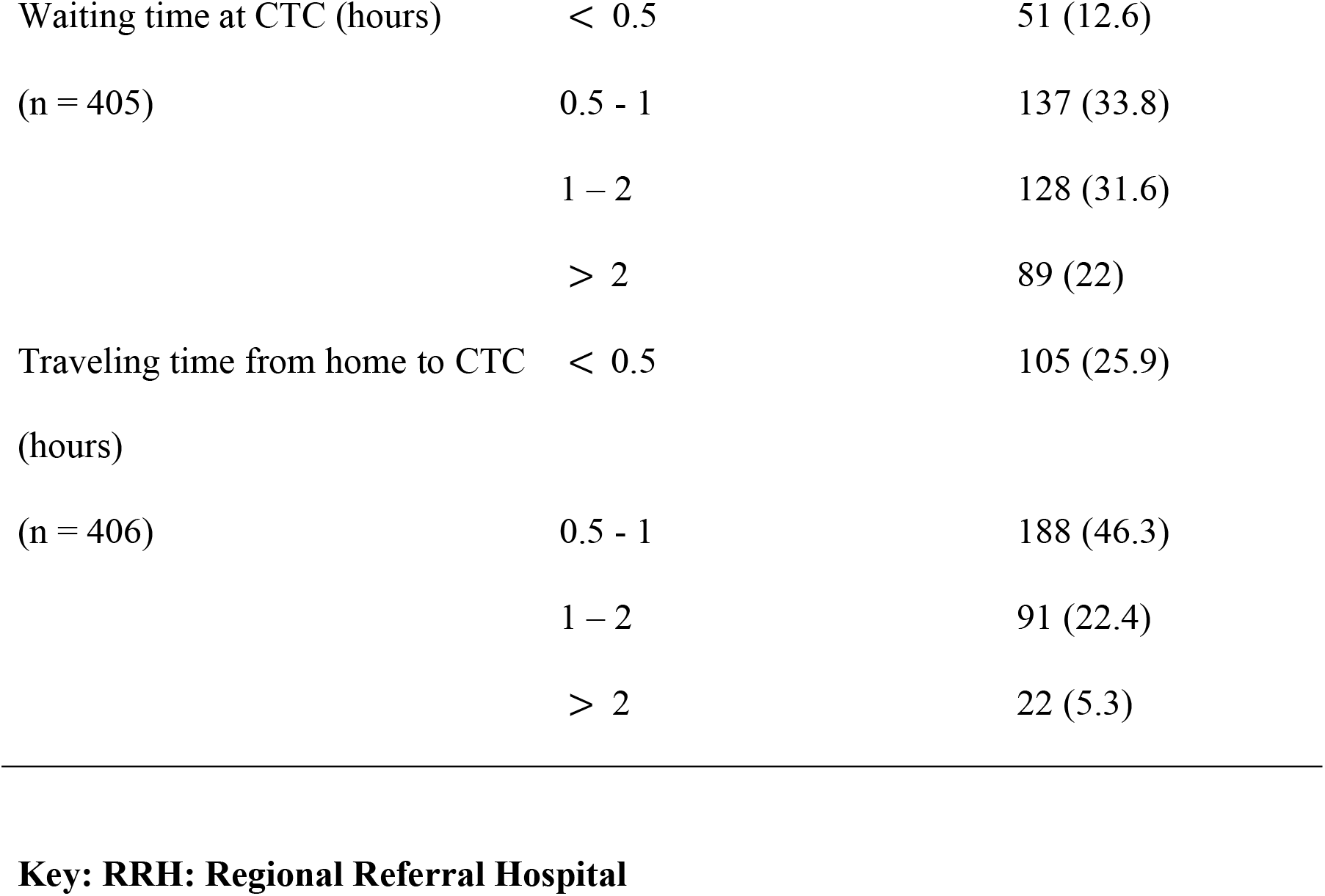
Participants’ socio-demographic characteristics.

| Variable | Category | Frequency N (%) |
| --- | --- | --- |
| Gender<br>(n = 406) | Female | 298 (73.2) |
|  | Male | 108 (26.5) |
| Age<br>(n = 404) | 18 - 24 | 33 (8.2) |
|  | 25 - 44 | 202 (50.0) |
|  | 45 - 54 | 108 (26.7) |
|  | 55 - 64 | 48 (11.9) |
|  | ≥ 65 | 13 (3.2) |
| Hospital facility<br>(n = 406) | Amana RRH | 205 (50.4) |
|  | Temeke RRH | 201 (49.4) |
| Start ARV use (years) | < 2 | 80 (20.3) |
| (n = 395) |  |  |
|  | 3 - 6 | 103 (26.1) |
|  | 7 - 12 | 151 (38.2) |
|  | ≥ 12 | 61 (15.4) |
| Marital status | Married | 171 (42.0) |
| (n = 402) |  |  |
|  | Single | 106 (26.0) |
|  | Divorced | 79 (19.4) |
|  | Widowed | 46 (11.3) |
| On dolutegravir based regimen | Yes | 382 (94.6) |
| (n = 404) |  |  |
|  | No | 22 (5.4) |
| Refill interval (months) | 1 | 83 (20.4) |
| (n = 406) | 2 | 7 (1.7) |
|  | 3 | 215 (52.8) |
|  | 6 | 101 (24.8) |
| Cost for a single hospital visit | < 2000 | 283 (69.9) |
| (TZS) |  |  |
| (n = 405) |  |  |
|  | ≥ 2000 | 87 (21.4) |
|  | No cost | 35 (8.6) |
| Waiting time at CTC (hours) | < 0.5 | 51 (12.6) |
| (n = 405) | 0.5 - 1 | 137 (33.8) |
|  | 1 – 2 | 128 (31.6) |
|  | > 2 | 89 (22) |
| Traveling time from home to CTC | < 0.5 | 105 (25.9) |
| (hours) |  |  |
| (n = 406) | 0.5 - 1 | 188 (46.3) |
|  | 1 – 2 | 91 (22.4) |
|  | > 2 | 22 (5.3) |

### Status disclosure among people living with HIV

The majority of the respondents, 378 (93.1%), disclosed their HIV status. The majority, 352 (93.6%) of those disclosing their HIV status, were consequently encouraged to take medication. Most of the participants, 246 (61.2%), reported concealing their HIV status to themselves, while 212 (52.7%) reported not having a reminder person or tool to assist them in taking their medication on time. When asked about the reminder person or a reminder tool, the majority of the participants, 52.7%, reported a lack of reminders. In contrast, the remaining participants, 24.4%, said their partners remind them. In comparison, 19.7% are reminded by their family members, and 6.7% reported using a phone and other devices to remind them to take medication (Table 2).

**Table 2.**
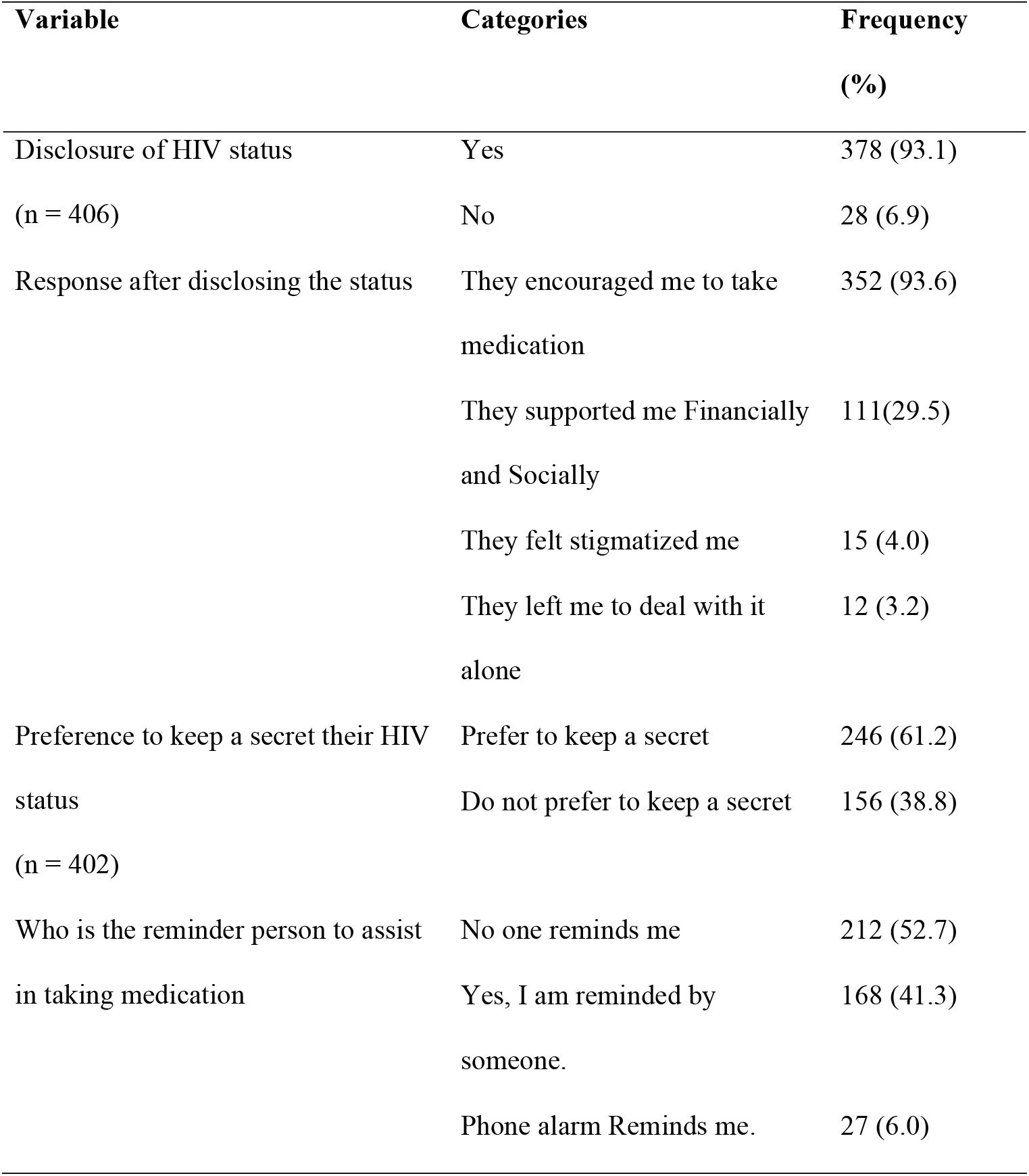
Status disclosure among people living with HIV.

### Self-discrimination and stigmatization

Of all participants, 355 (87.4%) reported that they preferred having a separate HIV clinic for HIV patients. When asked for a reason, most participants, 309 (87.0%), reported that a separate HIV CTC is a free space for HIV patients. Most participants, 275 (67.9%), agreed on having HIV hospital services if the HCW was a familiar person from their neighborhood (Table 3).

**Table 3.** Self-discrimination and stigmatization.

| Variable | Categories | Frequency (%) |
| --- | --- | --- |
| Preferred CTC location<br>(n = 399) | Having separated HIV clinic | 355 (87.4) |
|  | Inclusive HIV clinic | 26 (6.4) |
|  | Anyhow is better | 18 (4.4) |
|  | None of the above | 7 (1.7) |
| Why prefer separated HIV CTC | It is a free space for HIV patients | 309 (87.0) |
|  | I am comfortable seeing others with HIV, knowing that I am not alone | 99 (27.9) |
|  | I just like how the things are | 35 (9.9) |
|  | So as to avoid the long waiting queue at the inclusive clinic | 28 (7.9) |
| Would you have your HIV hospital services if<br>the health care worker is a person you know<br>from your neighborhood?<br>(n = 405) | Yes | 275 (67.9) |
|  | No | 130 (32.1) |
| Why not receive the hospital service from<br>familiar HCWs from the neighborhood | Because of the stigma that I will receive | 56 (43.4) |
|  | Because of fear that HCW will disclose my health status | 53 (41.1) |
|  | Because I do not prefer people who know me to know my health status | 30 (23.3) |
| Which container is used to store the medication | Inside its container | 320 (78.8) |
|  | Anywhere but not inside its container | 36 (8.9) |
|  | Inside my handbag | 34 (8.4) |
|  | Inside plastic bag | 30 (7.4) |
|  | Inside an envelope | 15 (3.7) |
|  | Inside of the pockets of my clothes | 10 (2.5) |
| To avoid the noise made by the containers, what do you add to the container | I am not bothered by the sound; I do not do anything | 227 (73.7) |
|  | I add a clean Handkerchief inside the container | 31 (10.1) |
|  | I add leaflet paper inside the containers | 27 (8.8) |
|  | I add clean cotton inside the container | 18 (5.8) |
|  | I only take my medication when I am at home. I do not move with them | 16 (5.2) |
|  | I do not move with medication inside the container. I keep them in the hospital plastic bags | 10 (3.2) |
| Where do you store ARV medication at home | On top of the cupboard | 123 (30.3) |
|  | Inside clothes cupboard | 118 (29.1) |
|  | Under the bed | 83 (20.4) |
|  | Inside backpack | 35 (8.6) |
|  | In between clothes in the suitcase | 29 (7.1) |
|  | Inside dishes cupboard | 19 (4.7) |
|  | Inside the refrigerator | 7 (1.7) |
|  | Near the window | 6 (1.5) |
|  | Inside a drawer | 5 (1.2) |
|  | In the car | 3 (0.7) |

### Factors associated with viral load suppression

Analysis of factors associated with viral load suppression found that viruses were more suppressed among female patients than their male counterparts (p = 0.036). Those on the dolutegravir-based regimen also suppressed viral load more than those on other regimens (p = 0.050). Participants with a refill interval of three months had more viral suppression than those with six months and additional months of refill interval with a p-value < 0.000 (p = 0.004) (Table 4).

**Table 4.**
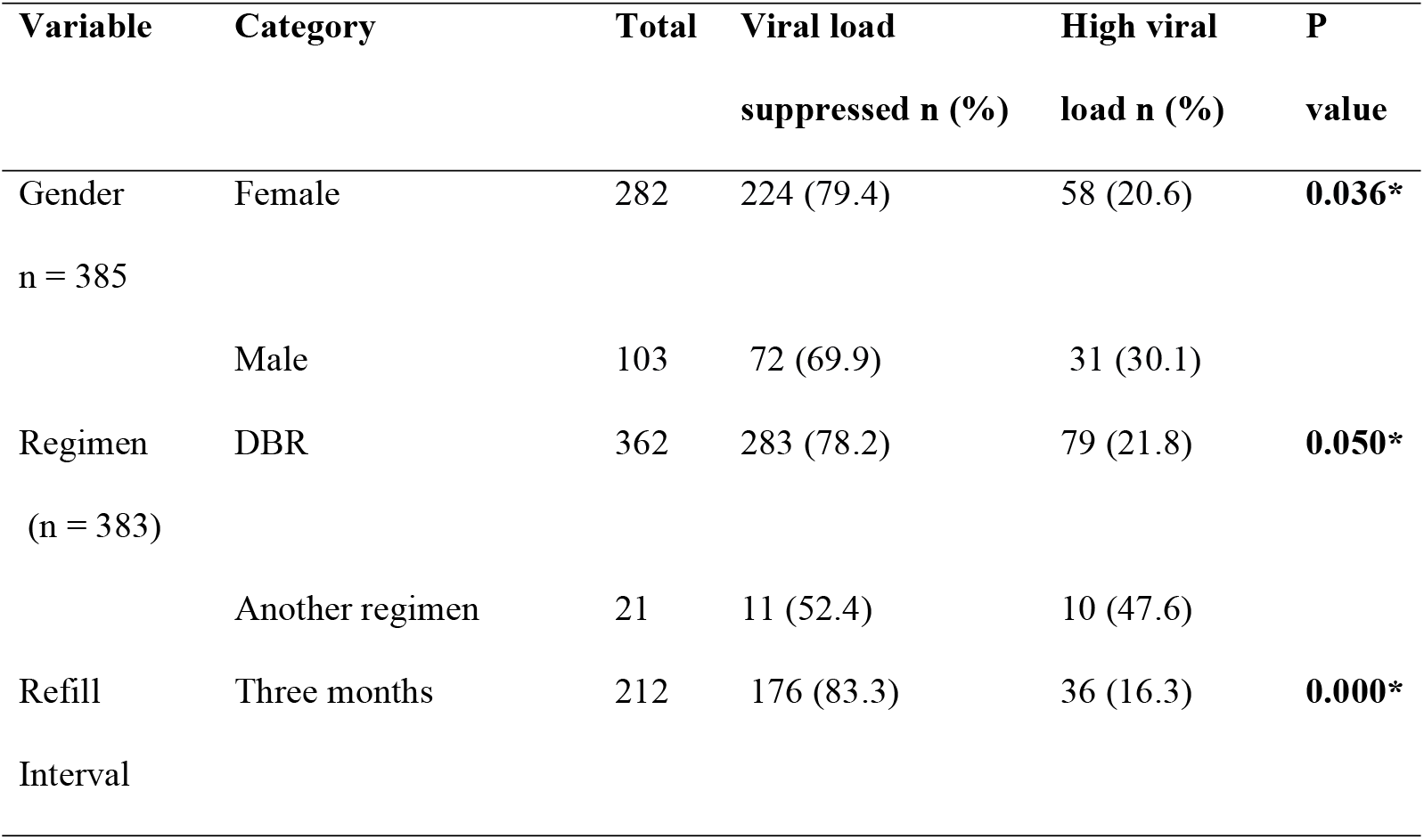
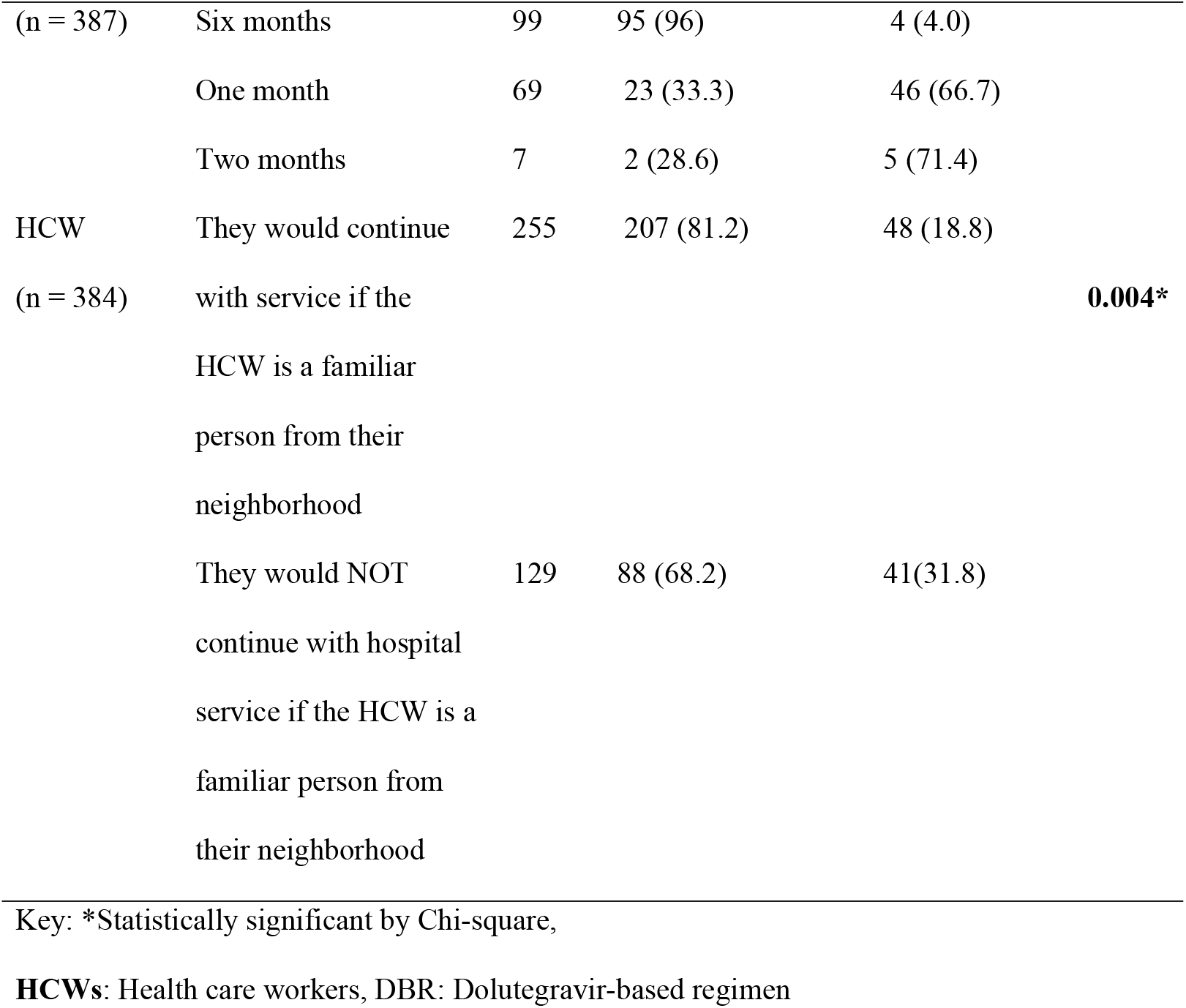
Factors associated with viral load suppression.

### Regression analysis of factors associated with viral load suppression

On a multivariate analysis, male patients were 60% less likely to have viral load suppression compared to female patients (adjusted odds ratio [aOR]: 0.4, 95% confidence interval [95%]: 0.19 – 0.77, p = 0.007). Refill interval was significantly associated with viral load suppression. For example, patients with one-month refill intervals had odds of 0.01 (95%CI: 0.003-0.42, p < 0.0001) as compared to those with six months refill intervals (Table 5).

**Table 5.** Multinomial logistics regression of factors associated with viral load suppression.

| Variable | Category | aOR (95%CI) | P value |
| --- | --- | --- | --- |
| Gender | Male | 0.4 (0.19 - 0.77) | 0.007* |
|  | Female | Reference | < 0.0001* |
| Refill interval | One month | 0.01 (0.003 – 0.42) | < 0.0001* |
|  | Two months | 0.01 (0.00 - 0.088) | < 0.0001* |
|  | Three months | 0.2 (0.054 - 0.566) | 0.004* |
|  | Six months | Reference |  |
| Adherence | Adherent | 1.2 (0.552 - 2.408) | 0.705 |
|  | Non-adherent | Reference |  |
| Name of facility | Temeke RRH | 1.5 (0.735 – 3.02) | 0.269 |
|  | Amana RRH | Reference |  |
| Regimen used | DBR | 1.1 (0.125 – 9.384) | 0.942 |
|  | Non-DBR | Reference |  |

\*Statistically significant, **aOR**: Adjusted Odds Ratio, **95%CI**: 95% confidence interval,
**RRH**: Regional referral hospital, **DBR**: Dolutegravir-based regimen

## Discussion

The study was conducted one year after introducing the dolutegravir-based regimen as the preferred default first line for managing HIV among adults in Tanzania [15], making the study the first to report the proportion of people who have been switched to dolutegravir-based regimens. This study found that most participants (87.4 %) preferred having a separate HIV clinic for HIV patients. When asked for a reason, they responded that a separate clinic would ensure their privacy by concealing their daily medication to the community surrounding PLHIV. It was further revealed that PLHIV tends to change the containers of medication. Most PLHIV reported using plastic bags, envelopes and handbags to carry their medication. This practice impairs the quality of ARV and, in turn, may result in low viral load suppression. However, in our study, the variable did not significantly affect viral load suppression.

When asked where they store their medication at home, most study participants (30.3%) responded that they hide their medication in the cupboard. This was consistent with a study done in South Africa by Flynn and colleagues [17], which indicated that most participants reported using the refrigerator or a typical cupboard to store their medication [17].

In addition, our study revealed that 78.2% of the respondents on dolutegravir-based regimens achieved viral load suppression. This proportion is less compared to the study done in Uganda in 2018, where 94% of PLHIV on dolutegravir achieved viral load suppression [5]. Also, this finding was below national data on viral load suppression, where 87.0% of adults on ART in Tanzania were reported to have suppressed viral loads [15]. However, this finding may be limited to a period of one year after the introduction of the dolultegravir regimen in Tanzania.

The female gender was significantly associated with viral load suppression. This is inconsistent with other studies in Uganda in 2014-2015 on factors associated with viral load non-suppression [18]. This study’s significant association between viral load suppression and refill time interval may be because the Tanzania National treatment guideline prescribes that PLHIV who consistently maintain viral load suppression are given a longer refill time interval [15].

Most of the study participants (67.9%) agreed on having HIV hospital services if the HCW is a person they know from their neighborhood compared to the 32.1% of participants who did not agree to be served by familiar healthcare personnel. Of all PLHIV not willing to be served by a familiar HCW, 43.4% reported the fear of stigmatization, while 41.1% feared that HCW would disclose their health status. Similar results were reported in the perpetuation of stigma by HCWs may have a more significant impact on the continuum of care outcomes of PLWH in research [19]. There was a significant association between viral load suppression and participants’ HCW preference for getting hospital service from the HCW familiar to them and coming from their neighborhood. The participants who were not particular about the HCW who served them were more likely to have viral suppression compared to those who would not continue with hospital service if the HCW was a familiar person from their neighborhood [20].

Our results are based on the self-reported perception of stigma. Self-reported data is usually subject to potential recall bias and social desirability. To minimize this bias, some information, such as socio-demographic characteristics and viral load information, was obtained from hospital records. Also, patients were requested to skip questions they did not remember rather than guessing the responses.

## Conclusion

Elements of self-perceived stigma appeared to influence the viral load suppression among PLHIV in Dar es Salaam, Tanzania. Factors such as gender and refill time interval were significantly associated with viral load suppression among HIV patients in the Dar es Salaam region. More counseling on stigmatization and its effects on PLHIV is needed to stop self-stigmatization.

## Data Availability

All data set used to draw this conclusions are included in the main paper

## Acknowledgments

We thank all the participants who agreed and consented to participate in this study. We also thank health care providers at Centres for Treatment and Counselling for their dedicated cooperation during the data collection phase.

## References

1. Bvochora T, Satyanarayana S, Takarinda KC, Bara H, Chonzi P, Komtenza B, et al. Enhanced adherence counselling and viral load suppression in HIV seropositive patients with an initial high viral load in Harare, Zimbabwe: Operational issues. Mor O, editor. PLoS One. 2019;14: e0211326. doi:10.1371/journal.pone.0211326

2. Omosanya OE, Elegbede OT, Agboola SM, Isinkaye AO, Omopariola OA. Effects of Stigmatization/Discrimination on Antiretroviral Therapy Adherence among HIV-Infected Patients in a Rural Tertiary Medical Center in Nigeria. J Int Assoc Provid AIDS Care. 2014;13: 260–263. doi:10.1177/2325957413475482

3. Bernays S, Paparini S, Seeley J, Rhodes T. “Not Taking it Will Just be Like a Sin”: Young People Living with HIV and the Stigmatization of Less-Than-Perfect Adherence to Antiretroviral Therapy. Med Anthropol. 2017;36: 485–499. doi:10.1080/01459740.2017.1306856

4. Tsai AC, Bangsberg DR, Kegeles SM, Katz IT, Haberer JE, Muzoora C, et al. Internalized Stigma, Social Distance, and Disclosure of HIV Seropositivity in Rural Uganda. Ann Behav Med. 2013;46: 285–294. doi:10.1007/s12160-013-9514-6

5. Nabitaka VM, Nawaggi P, Campbell J, Conroy J, Harwell J, Magambo K, et al. High acceptability and viral suppression of patients on Dolutegravir-based first-line regimens in pilot sites in Uganda: A mixed-methods prospective cohort study. Torpey K, editor. PLoS One. 2020;15: e0232419. doi:10.1371/journal.pone.0232419

6. Dean Wantland W-TC. Engagement with Health Care Providers Affects Self-Efficacy, Self-Esteem, Medication Adherence and Quality of Life in People Living with HIV. J AIDS Clin Res. 2013;04: 1–14. doi:10.4172/2155-6113.1000256

7. United Republic of Tanzania. National Beaural of Statistics: 2012 Population and Housing Census Population Distribution by Administrative areas. Natl Bur Stat Minist Financ. 2013; 177,180.

8. Pourhoseingholi MA, Vahedi M, Rahimzadeh M. Sample size calculation in medical studies. Gastroenterol Hepatol from bed to bench. 2013;6: 14–7.

9. Sangeda RZ, Mosha F, Aboud S, Kamuhabwa A, Chalamilla G, Vercauteren J, et al. Predictors of non adherence to antiretroviral therapy at an urban HIV care and treatment center in Tanzania. Drug Healthc Patient Saf. 2018;10: 79–88. doi:10.2147/DHPS.S143178

10. Dehens J, de Hemptinne M, Galouchka M, Sajud A, van Otzel RP, Vanhoorebeeck C, et al. Exploring the value and acceptability of peer support in the process of improving adherence to HIV antiretroviral drugs in Tanzania, Dar-es-Salaam. Transdiscipl Insights. 2017;1: 9–32. doi:10.11116/TDI2017.1.2

11. Mosha F, Ledwaba J, Ndugulile F, Ng’ang’a Z, Nsubuga P, Morris L, et al. Clinical and virological response to antiretroviral drugs among HIV patients on first-line treatment in Dar-es-Salaam, Tanzania. J Infect Dev Ctries. 2014;8: 845–52. doi:10.3855/jidc.3879

12. Harris PA, Taylor R, Minor BL, Elliott V, Fernandez M, O’Neal L, et al. The REDCap consortium: Building an international community of software platform partners. J Biomed Inform. 2019;95: 103208. doi:10.1016/j.jbi.2019.103208

13. Harris PA, Taylor R, Thielke R, Payne J, Gonzalez N, Conde JG. Research electronic data capture (REDCap)--a metadata-driven methodology and workflow process for providing translational research informatics support. J Biomed Inform. 2009;42: 377–81. doi:10.1016/j.jbi.2008.08.010

14. USAID. Understanding and Challenging HIV Stigma toward Entertainment Workers. 2007 [cited 19 Aug 2021] p. 143. Available: https://www.icrw.org/wp-content/uploads/2016/10/Understanding-and-Challenging-HIV-Stigma-toward-Entertainment-Workers-Toolkit-for-Action.pdf

15. United Republic of Tanzania. National Guidelines for the management of HIV and AIDS in Tanzania. 2019 [cited 30 May 2021]. Available: http://www.nacp.go.tz/download/national-guidelines-for-the-management-of-hiv-and-aids/

16. Kilipamwambu A, Bwire GM, Myemba DT, Njiro BJ, Majigo M V. WHO/INRUD core prescribing indicators and antibiotic utilization patterns among primary health care facilities in Ilala district, Tanzania. JAC-Antimicrobial Resist. 2021;3: 1–7. doi:10.1093/jacamr/dlab049

17. Flynn AD, Scheuerle RL, Galgon G, Gerrard SE, Netshandama VO. An assessment of infant medication administration and storage practices in selected communities in the Vhembe District of Limpopo Province, South Africa. Heal SA Gesondheid. 2019;24: 1–7. doi:10.4102/hsag.v24i0.1075

18. Bulage L, Ssewanyana I, Nankabirwa V, Nsubuga F, Kihembo C, Pande G, et al. Factors Associated with Virological Non-suppression among HIV-Positive Patients on Antiretroviral Therapy in Uganda, August 2014–July 2015. BMC Infect Dis. 2017;17: 326. doi:10.1186/s12879-017-2428-3

19. Algarin AB, Sheehan DM, Varas-Diaz N, Fennie KP, Zhou Z, Spencer EC, et al. Health Care-Specific Enacted HIV-Related Stigma’s Association with Antiretroviral Therapy Adherence and Viral Suppression Among People Living with HIV in Florida. AIDS Patient Care STDS. 2020;34: 316–326. doi:10.1089/apc.2020.0031

20. Kiekens A, Dehens J, de Hemptinne M, Galouchka M, Vanhoorebeeck C, van Otzel RP, et al. HIV-related Peer Support in Dar es Salaam: A Pilot Questionnaire Inquiry. Transdiscipl Insights. 2019;3: 1–18. doi:10.11116/TDI2019.3.1

